# Distinct systemic and mucosal immune responses to SARS-CoV-2

**DOI:** 10.1101/2021.03.01.21251633

**Authors:** Nikaïa Smith, Pedro Goncalves, Bruno Charbit, Ludivine Grzelak, Maxime Beretta, Cyril Planchais, Timothée Bruel, Vincent Rouilly, Vincent Bondet, Jérôme Hadjadj, Nader Yatim, Helene Pere, Sarah H Merkling, Solen Kernéis, Frederic Rieux-Laucat, Benjamin Terrier, Olivier Schwartz, Hugo Mouquet, Darragh Duffy, James P. Di Santo

## Abstract

Coordinated local mucosal and systemic immune responses following SARS-CoV-2 infection protect against COVID-19 pathologies or fail leading to severe clinical outcomes. To understand this process, we performed an integrated analysis of SARS-CoV-2 spike-specific antibodies, cytokines, viral load and 16S bacterial communities in paired nasopharyngeal swabs and plasma samples from a cohort of clinically distinct COVID-19 patients during acute infection. Plasma viral load was associated with systemic inflammatory cytokines that were elevated in severe COVID-19, and also with spike-specific neutralizing antibodies. In contrast, nasopharyngeal viral load correlated with SARS-CoV-2 humoral responses but inversely with interferon responses, the latter associating with protective microbial communities. Potential pathogenic microrganisms, often implicated in secondary respiratory infections, were associated with mucosal inflammation and elevated in severe COVID-19. Our results demonstrate distinct tissue compartmentalization of SARS-CoV-2 immune responses and highlight a role for the nasopharyngeal microbiome in regulating local and systemic immunity that determines COVID-19 clinical outcomes.

## Introduction

While SARS-CoV-2 (severe acute respiratory syndrome coronavirus 2) infection is responsible for COVID-19 (coronavirus disease 2019), the regulatory mechanisms underlying disease pathophysiology remain enigmatic. Clinical manifestations following SARS-CoV-2 infection are highly variable, ranging from asymptomatic or mild symptoms to severe pneumonia that can progress to acute respiratory distress syndrome (ARDS) (Huang et al., 2020b). It is still unclear whether disease progression is related to the viral infection itself, to the host immune response, to host co-morbidities or to a combination of these different factors (Williamson et al., 2020). Biomarkers to distinguish disease progression in COVID-19 include IL-6, CRP, d dimers, and LDH, yet our understanding of their role in disease pathophysiology remains limited (Fajgenbaum and June, 2020).

Analysis of immune responses in COVID-19 patients showed that SARS-CoV-2 suppresses activation of the innate immune system, including dendritic cells (Zhou et al., 2020a) and dampens antiviral type I and type III interferon responses (Blanco-Melo et al., 2020; Hadjadj et al., 2020), leading to an excessive proinflammatory macrophage activation. Despite overall peripheral lymphopenia, COVID-19 patients mount efficient SARS-CoV-2-specific memory T and B cell responses (Mathew et al., 2020). In particular, COVID-19 patients show increased numbers of plasma cells and generate specific neutralizing antibodies against the SARS-CoV-2 spike protein (Long et al., 2020; Rogers et al., 2020). Virus-specific T cell responses in the blood increase with disease severity suggesting that defects in adaptive immunity are not causal during early stages (Grifoni et al., 2020).

One severe clinical manifestation in COVID-19 patients is an extensive systemic immune reaction triggered by the excessive production of inflammatory mediators such as Monocyte Chemoattractant Protein-1 (MCP-1/CCL2), Macrophage Inflammatory Protein 1alpha (MIP1A/CCL3), IL-6, TNF-α and IL-10 (Merad and Martin, 2020). SARS-CoV-2-associated hyperinflammation can promote a pathological hypercoagulable state with increased mortality for COVID-19 patients. The systemic hyperinflammation correlates with peripheral SARS-CoV-2 RNA loads suggesting that it represents a form of ‘viral’ sepsis (Li et al., 2020). Still, the exact mechanism underlying this phenomenon remains to be determined.

Upon initial exposure, SARS-CoV-2 is thought to infect hACE2-expressing epithelial cells in the upper respiratory tract (Sungnak et al., 2020). At this stage, early defense mechanisms likely limit viral replication in most individuals and prevent further disease progression. These may include physio-chemical barriers (mucus, metabolites), as well as innate immune defense proteins (cytokines, interferons) that are constitutively produced or induced upon infection. Adaptive immune mechanisms, including secretory IgA, play a critical role in barrier function at mucosal sites. In the context of SARS-CoV-2 infection, several studies have documented presence of virus-specific IgG and IgA in blood, saliva and nasopharyngeal samples of COVID-19 patients (Cervia et al., 2020; Isho et al., 2020; Sterlin et al., 2020; Zohar et al., 2020). Still, how local and systemic immunity following SARS-CoV-2 infection is established and the factors that regulate this process are poorly understood.

Here we applied an integrated systems approach to identify the factors that regulate local and systemic immunity to SARS-CoV-2 using a cohort of COVID-19 patients with varying clinical severity. Our results reveal distinct responses between nasal and systemic immunity, with a strong impact on the nasal cytokine response and microbiome in severe COVID-19 disease. These results may support new strategies for management of patients infected with SARS-CoV-2.

## Results

### Systemic and mucosal antibody responses in COVID-19 patients

Little is known about how systemic and nasopharyngeal humoral immune responses are coordinated during SARS-CoV-2 infection. To better understand the regulatory mechanisms controlling SARS-CoV-2-specific immune responses at the initial site of infection as well as systemically, we first measured Spike-specific IgG and IgA and total Ig in paired plasma and nasopharyngeal samples from hospitalized COVID-19 patients and healthy controls. The COVID-19 patient cohort consisted of PCR confirmed patients at 8-12 days post-symptom onset with distinct clinical classification (indicated here as moderate, severe and critical; (Hadjadj et al., 2020); see STAR Methods) as well as non-COVID-19 controls. To assess SARS-CoV-2-specific antibody responses, we used two complementary and sensitive assays to measure Spike-specific IgG and IgA: an ELISA-based approach using soluble trimeric CoV-2 spike protein and the ‘S-flow’ FACS-based approach using cell lines stably expressing surface CoV-2 spike (see STAR Methods for more details; (Fafi-Kremer et al., 2020; Grzelak et al., 2020). In line with previous reports (Long et al., 2020; Röltgen et al., 2020), we detected spike-specific IgG and IgA antibodies in plasma of COVID-19 patients (n= 49) but not in healthy controls with an increasing frequency and intensity dependent on disease severity (**Fig. 1A-C**). Robust systemic IgG and IgA responses were detected by ELISA and ‘S-flow’ approaches with excellent correlations between assays (**Supplemental Fig. 1A**) confirming previous reports (Grzelak et al., 2020). We next assessed the neutralization activity of plasma samples against SARS-CoV-2 using a pseudovirus infection assay (see STAR Methods (Grzelak et al., 2020). Neutralization capacity was clearly induced following SARS-CoV-2 infection and increased with clinical severity (**Fig. 1D-E**). Moreover, neutralization intensity was highly correlated with frequency of Spike-specific IgG and IgA (**Fig. 1F**). We did not find significant differences in plasma total IgM, IgG and IgA levels or in IgG subclass levels between healthy individuals and COVID-19 patients (**Supplemental Fig. 1B**).

**Fig. 1.**
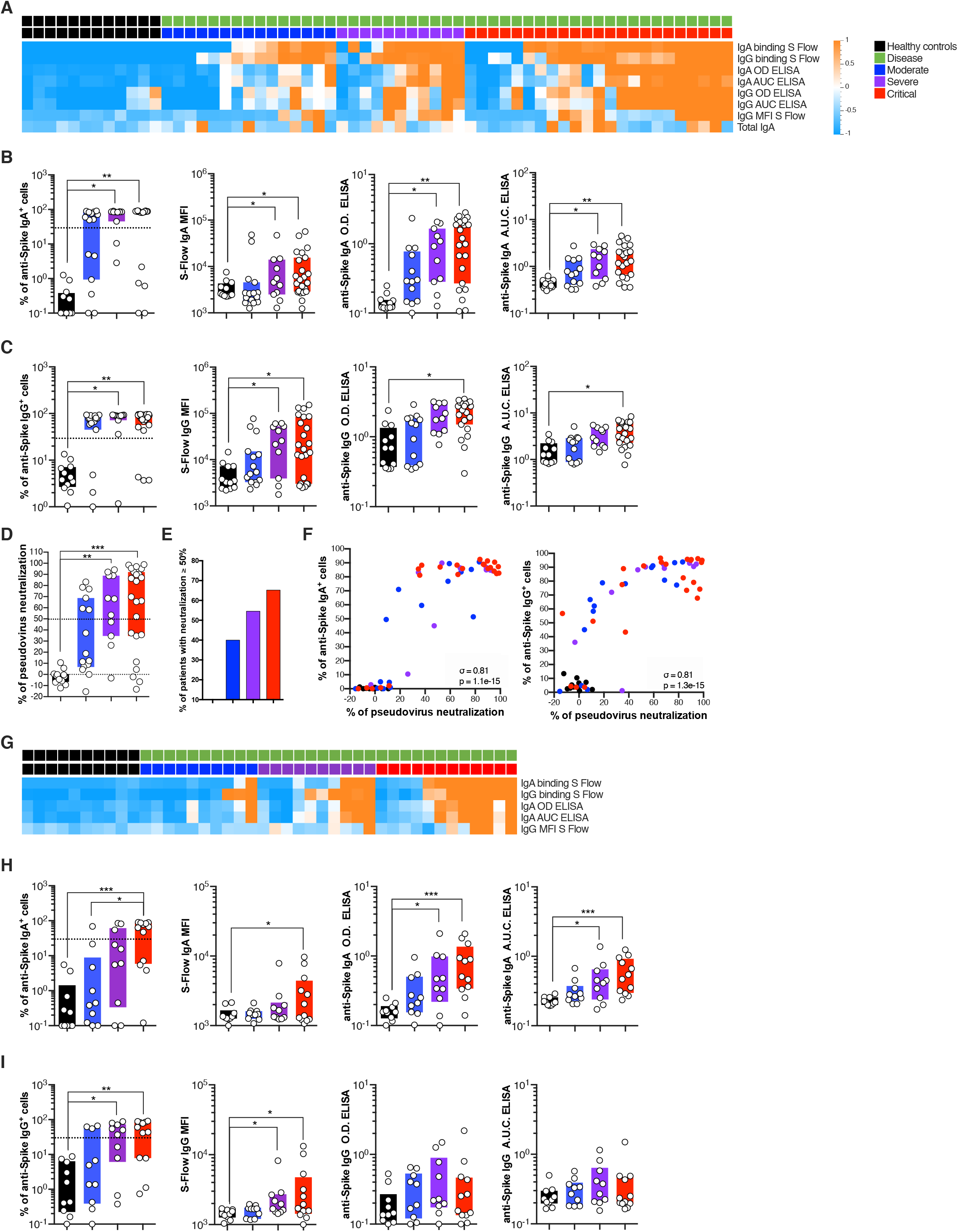
Systemic and mucosal antibody responses in COVID-19 patients. Antibodies were measured in the plasma (panels **A, B, C**) of healthy controls (n = 12 donors), mild to moderate (n = 15 patients), severe (n = 11 patients) and critical (n = 23 patients) or in the nasopharyngeal compartment (panels **G, H, I**) of healthy controls (n = 10 donors), mild to moderate (n = 10 patients), severe (n = 10 patients) and critical (n = 12 patients) using an ELISA-based approach using soluble CoV-2 spike protein (OD and AUC ‘ELISA’) and the ‘S-flow’ FACS-based approach using cell lines stably expressing surface CoV-2 spike (‘S_Flow’), Heatmap representation of statistically different (P<0.05) antibody responses between healthy controls and COVID-19 patients (moderate, severe, critical) in (**A**) plasma and (**G**) nasopharyngeal compartment. (**B-C**) and (**H-I**) Individual antibodies responses by patient severity. (**D**) Pseudovirus neutralization (%) against the SARS-CoV-2 spike protein was measured by analyzing luciferase-expressing pseudotypes. (**E**) Graph represents the percentage of patients with pseudotype neutralization above 50%. (**F**) Correlation plots between the pseudotype neutralization (%) and presence of anti-Spike IgA or IgG as measured by S_flow. In (**A**) and (**G**), P values were determined with the Mann-Whitney test between healthy and infected cases. In (**B, C, D, H** and **I**), box plots with median ± minimum to maximum. P values were determined with the Kruskal-Wallis test followed by with Dunn’s post-test for multiple group comparisons. In (**F**), σ represents Spearman coefficient and p the p value; *P < 0.05; **P < 0.01; ***P < 0.001.

We applied the same antibody assays to nasopharyngeal samples (n= 42) collected at the same time as the plasma from a majority of patients in this COVID-19 cohort. As with plasma, we found significantly increased frequency and intensity of spike-specific IgG and IgA responses in nasopharyngeal secretions as disease severity increased (**Fig. 1G-I**). Strong correlations between different spike-specific antibody assays in nasal samples were also observed (**Supplemental Fig. 1C)**. Interestingly, nasopharyngeal total IgA (but not total IgM or IgG or IgG subclass) levels were significantly elevated in critical COVID-19 patients (**Supplemental Fig. 1D)**. These results confirm and extend previous reports of robust local and systemic humoral responses against the SARS-CoV-2 spike protein in acute COVID-19 infection (Cervia et al., 2020).

### Heterogeneity of antibody responses in COVID-19 patients

As the majority of the nasopharyngeal samples had a paired plasma sample (n= 41), we next explored the relationship between local mucosal and systemic spike-specific antibody production in COVID-19 patients. This analysis confirmed previous reports (Cervia et al., 2020) but also revealed several unexpected patterns of anti-SARS-CoV-2 humoral immunity. First, the majority (88%) of COVID-19 patients seroconverted with spike-specific antibodies in their blood, which appeared to be independent of disease severity (**Fig. 2A**). Both spike-specific IgG and IgA were present in the majority of these seropositive COVID-19 individuals (**Fig. 2B; Supplemental Fig. 2A**). Second, overall ‘nasoconversion’ (presence of spike-specific IgG or IgA in nasopharyngeal secretions) was significantly less frequent than that observed for seroconversion (**Fig. 2A**). Nevertheless, in these ‘nasoconverters’, spike-specific IgG and IgA were still largely co-detected (**Fig. 2B; Supplemental Fig. 2A**). Finally, a small fraction of COVID-19 patients (12.25%; 6/49) did not show IgG or IgA seroconversion (**Fig. 2A**), despite having been exposed to SARS-CoV-2 (all COVID-19 patients were confirmed PCR positive). Together, these results suggest a complex patient-specific control of local mucosal and systemic antibody responses at this early time point (day 8-12) following SARS-CoV-2 infection.

**Fig. 2.**
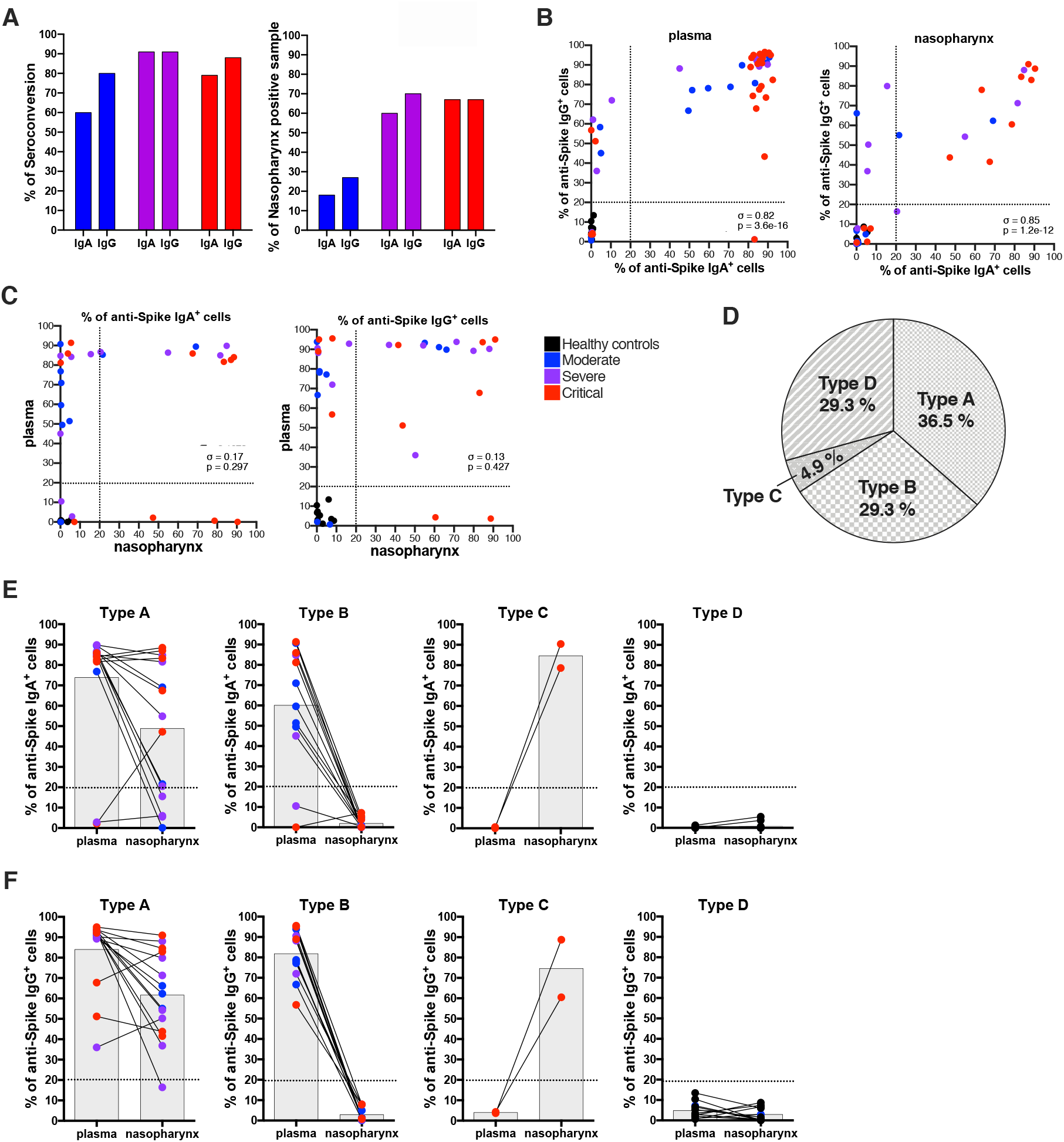
Heterogeneity of antibody responses in COVID-19 patients. IgA and IgG were assessed by S_flow using cell lines stably expressing surface CoV-2 spike. (**A**) IgA and IgG seroconversion (%) in plasma and ‘nasoconversion’ (% of nasopharynx positive samples) versus disease severity. (**B**) Correlation plots between the anti-Spike IgA+ and IgG+ cells (%) in plasma and in nasopharynx. (**C**) Correlation plots between plasma and nasopharynx anti-Spike antibody responses. (**D**) Representation of antibody conversion among the patients; type ‘A’: naso-positive and sero-positive patients; type ‘B’: naso-negative and sero-positive patients; type ‘C’: naso-positive and sero-negative patients; type ‘D’: naso-negative and sero-negative patients. (**E**) Representation of anti-Spike antibody responses in the different compartment per patient type. In (**B**) and (**C**), σ represents Spearman coefficient and p the p value.

We next compared systemic and local mucosal spike-specific IgG and IgA responses in individual COVID-19 patients. Surprisingly, systemic (plasma) and local (nasopharynx) spike-specific responses within individuals were not correlated (**Fig. 2C**). This was apparent when comparing spike-specific IgG or IgA responses in plasma versus nasopharynx or when cross-comparing IgG with IgA responses (**Supplemental Fig. 2B-D**). This result suggests independent regulation of mucosal and systemic immune responses to SARS-CoV-2.

We next subclassified COVID-19 patients and controls for which blood and nasal samples were both available (n = 41) based on the presence or absence of spike-specific IgG and/or IgA in the plasma (P) or nasopharynx (N) as type ‘A’ (PN+; 29.3%), type ‘B’ (P+; 36.5%), type ‘C’ (N+; 4.9%) or type ‘D’ (PN-; 29.3%) responders (**Fig. 2D, E; Supplemental Fig. 2E**). As expected, all controls were type ‘D’ (sero-/naso-negative), but 2 moderate COVID-19 patients were sero- and naso-negative at this time point (**Fig. 1A, D**). Interestingly, 2 critical patients showed an absence of spike-specific antibodies in the plasma but strong spike-specific IgG and IgA responses in the nasopharynx (**Fig. 2D, E; Supplemental Fig. 2E**) identifying these patients as type ‘C’ responders. The remaining COVID-19 patients were split between type ‘A’ and type ‘B’ responders that were not significantly enriched for any particular disease severity (**Supplemental ig. 2E**). Taken together, these results demonstrate heterogeneous SARS-CoV-2 antibody responses within systemic and local mucosal sites that suggest tissue-dependent regulation of this process.

### Differential systemic and mucosal cytokine responses in COVID-19 patients

In order to better understand the mechanisms that could regulate mucosal and systemic spike-specific antibody responses to SARS-CoV-2, we measured the concentrations of 46 cytokines in plasma and nasopharyngeal samples. In plasma, 13 cytokines were significantly different (p<0.05, q<0.2, n=61 samples) between the healthy donors and COVID-19 patients regardless of disease severity (**Fig. 3A, B**). This included VEGF, FGF, IL-1RA, IL-6, TNF-α, IL-10, CCL2/MCP-1, CXCL10/IP-10, CCL3/MIP-1α, CCL19/MIP-3β, PD-L1, G-CSF and Granzyme B. In contrast, a strikingly different cytokine profile was observed in the nasopharynx: using the same significance cutoff (p<0.05, q<0.2, n=42 samples), a limited and largely non-overlapping set of 7 cytokines was found to be significantly different between the healthy donors and COVID-19 patients (**Fig. 3C, D**). This included IL-33, IFNα2, IFNλ3, IFNβ and IFNγ which were decreased in the nasopharynx of COVID-19 patients while IL-10 and CCL2/MCP-1 were increased as compared to healthy controls (**Fig. 3C, D)**. Of the two plasma and nasopharyngeal cytokines differentially expressed between healthy and COVID-19 patients (IL-10, CCL2/MCP-1), both were increased during infection in plasma and nasopharyngeal samples (**Fig. 3B, D)**. These results confirm and extend previous reports identifying enhanced inflammatory and diminished interferon responses in the context of SARS-CoV-2 infection (Bastard et al., 2020; Blanco-Melo et al., 2020; Hadjadj et al., 2020) but show that nasopharyngeal cytokine responses are regulated in a distinct fashion.

**Fig. 3.**
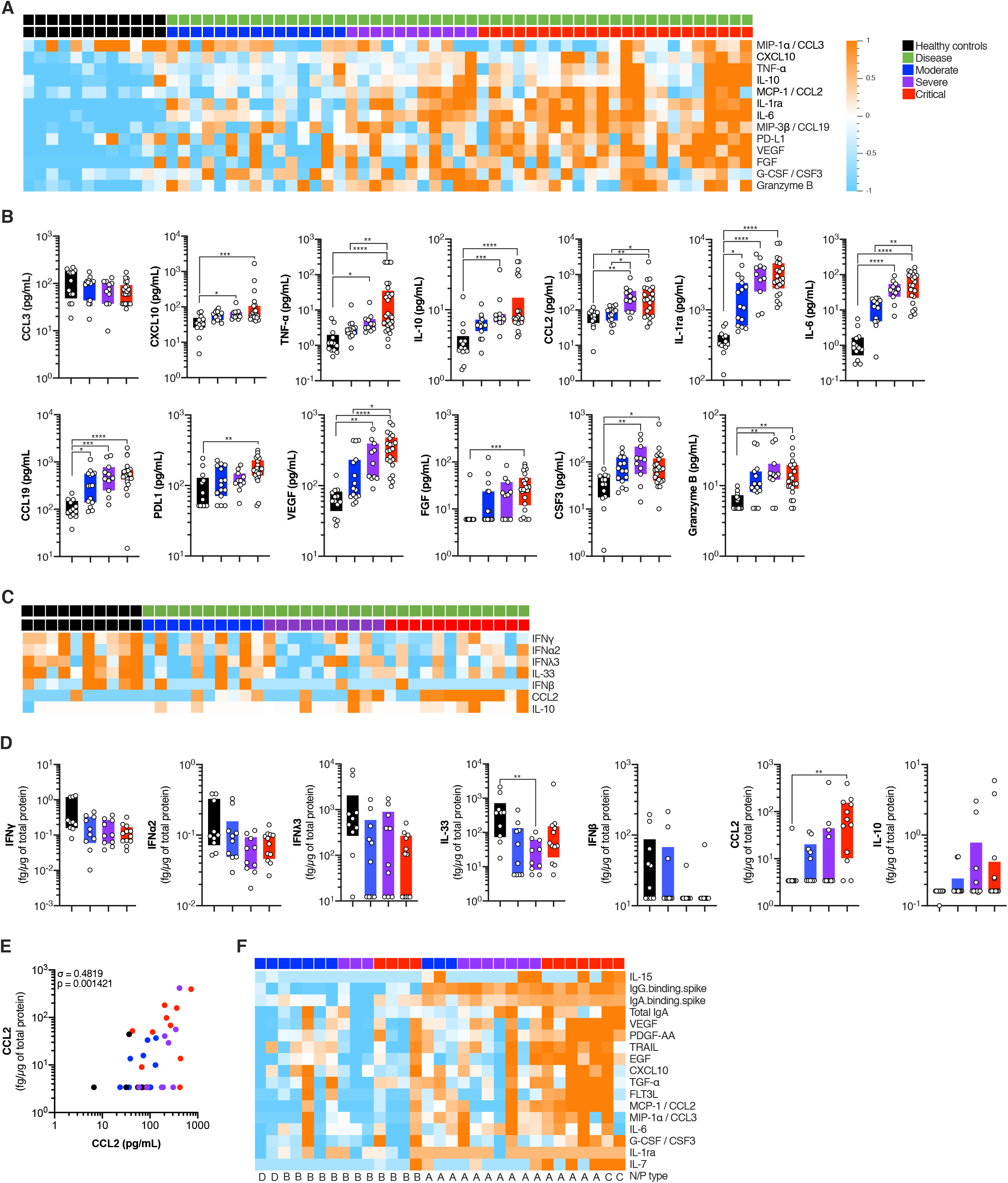
Systemic and mucosal cytokines production in COVID-19 patients. Cytokines were measured in the plasma (panels **A, B**) of healthy controls (n = 12 donors), mild to moderate (n = 15 patients), severe (n = 11 patients) and critical (n = 23 patients) or in the nasopharyngeal compartment (panels **C, D**) of healthy controls (n = 10 donors), mild to moderate (n = 10 patients), severe (n = 10 patients) and critical (n = 12 patients) using a bead-based multiplexed immunoassay system Luminex or the digital Elisa Simoa (IFNα, IFNβ, IFNγ, IL-6, IL-17A, IL-10, TNF-α). (**A**) and (**C**) Heatmap representation of statistically different cytokines (P<0.05) between healthy controls and COVID-19 patients (moderate, severe, critical), ordered by hierarchical clustering. Up-regulated cytokines are shown in orange, and down-regulated in blue. (**B**) and (**D**) Individual cytokine concentration plots by patient severity. In (**A**) and (**C**), P values were determined with the Mann-Whitney test between healthy and infected cases. In (**B**) and (**D**), box plots with median ± minimum to maximum. P values were determined with the Kruskal-Wallis test followed by Dunn’s post-test for multiple group comparisons; *P < 0.05; **P < 0.01; ***P < 0.001. (E) Heatmap representation of statistically different cytokines (P<0.05) in patients having nasopharyngeal spike-specific antibodies (type ‘A’ and ‘C’) compared with those lacking these antibodies (‘B’ and ‘D’). Up-regulated cytokines are shown in orange, and down-regulated cytokines are shown in blue.

### Nasopharyngeal and systemic cytokine responses stratify COVID-19 disease severity

Previous studies have reported perturbed systemic cytokine production as a hallmark of disease severity in COVID-19 patients (Lucas et al., 2020; Merad and Martin, 2020). Extending our previous work with this cohort (Hadjadj et al., 2020), we identified 10 circulating cytokines that were significantly different (p<0.05, q<0.2, n=49 samples) between the critical and non-critical (mild/moderate and severe) COVID-19 cases (**Supplemental Fig. 3A,B**). This included IL-6, IL-10, CCL20/MIP-3α, VEGF, FGF, PD-L1, TNF-α, IL-1β and IL-1RA which increased with disease severity, and IFNα2 which decreased with severity extending our previous results (Hadjadj et al., 2020).

We also studied whether nasopharyngeal cytokine profiles varied with disease severity. Using the same significance cutoff (p<0.05, q<0.2, n=32 samples), we found that 13 nasopharyngeal cytokines were differently regulated between the critical and non-critical COVID-19 cases (**Supplemental Fig. 3C, D**). Interestingly, only two cytokines (CCL2/MCP-1, VEGF) overlapped with the plasma dataset (**Fig. 3E**), whereas other nasal cytokines including FLT3-L, EGF, CXCL1/GROα, PDGF-AA, IL-7 and TGF-α were significantly increased with worsening disease severity (**Supplemental Fig. 3C,D**). Taken together, these results suggest that cytokine responses are compartmentalized during SARS-CoV-2 infection and are regulated, similar to spike-specific antibodies, in a tissue-dependent fashion.

As certain cytokines are known to negatively regulate antibody responses (ie type I interferons; Hensley et al., 2007; Moseman et al., 2016), we performed hierarchical clustering of plasma and nasopharyngeal cytokines to identify possible associations that may explain the distinct spike-specific humoral responses (**Fig. 2D, E**). Analysis of nasopharyngeal cytokines showed higher levels of IL-15, VEGF, PDGF-AA, TRAIL, EGF, CXCL10, TGF-a, FLT3L, CCL2, CCL3, IL-6, G-CSF, IL-1RA and IL-7 in nasotypes ‘A’ and ‘C’ (with nasal spike-specific Ab) compared to nasotypes ‘B’ and ‘D’ (without nasal spike-specific Ab) (**Fig. 3F**) suggesting that inflammation could be involved in local antibody generation. Notably, the interferon response showed no obvious associations with the presence or absence of viral-specific antibodies. These results provide further evidence for distinct host immune responses to SARS-CoV-2 infection at local and systemic levels.

### Viral load drives differential systemic and mucosal immune responses

We next asked the question of whether the virus may be directly influencing this tissue specific immunity. We correlated spike-specific antibody and cytokine responses with viral load as measured in nasopharynx by RT-PCR, and in plasma with a droplet digital PCR assay as previously described (see STAR Methods, (Veyer et al., 2020). We found that viral load was increased in both local mucosal as well as systemic compartments in COVID-19 patients and were partially correlated (**Fig. 4A, B**). Interestingly, while plasma viral load increased with increasing disease severity, nasopharyngeal viral load was largely independent of the clinical presentation (**Fig. 4A**) consistent with previous reports (Chen et al., 2020; Fajnzylber et al., 2020; Zou et al., 2020). In order to gain insight into how viral load may influence immune responses, we performed multidimensional scaling (MDS) which is a way of visualizing the level of similarity of individual cases of a dataset (in this case viral load, cytokines and antibody response characteristics). From the MDS projection and correlation matrix of our plasma dataset (**Fig. 4C; Supplemental Fig. 4A**), we could see that viral load was positively associated with the systemic inflammatory response (IL-6, TNFα, MIP-3β) and several regulatory cytokines (IL-10, IL-1RA) but not with the anti-viral interferon response (IFNα2) (**Fig. 4C, D**). These results are in line with several reports of SARS-CoV-2-dependent induction of hyper-inflammation as well as the critical role for interferon responses in controlling initial infection (Chen et al., 2020; Fajnzylber et al., 2020; Hadjadj et al., 2020). Interestingly, plasma viral load was positioned distinctly from pseudo-neutralization activity (that formed a tight cluster with systemic spike-specific IgG and IgA) (**Fig. 4C**). Nevertheless, viral load was positively correlated with these virus-specific antibody responses (**Fig. 4D; Supplemental Fig. 4A**) indicating a role for viral load in driving spike-specific humoral immunity.

**Fig. 4.**
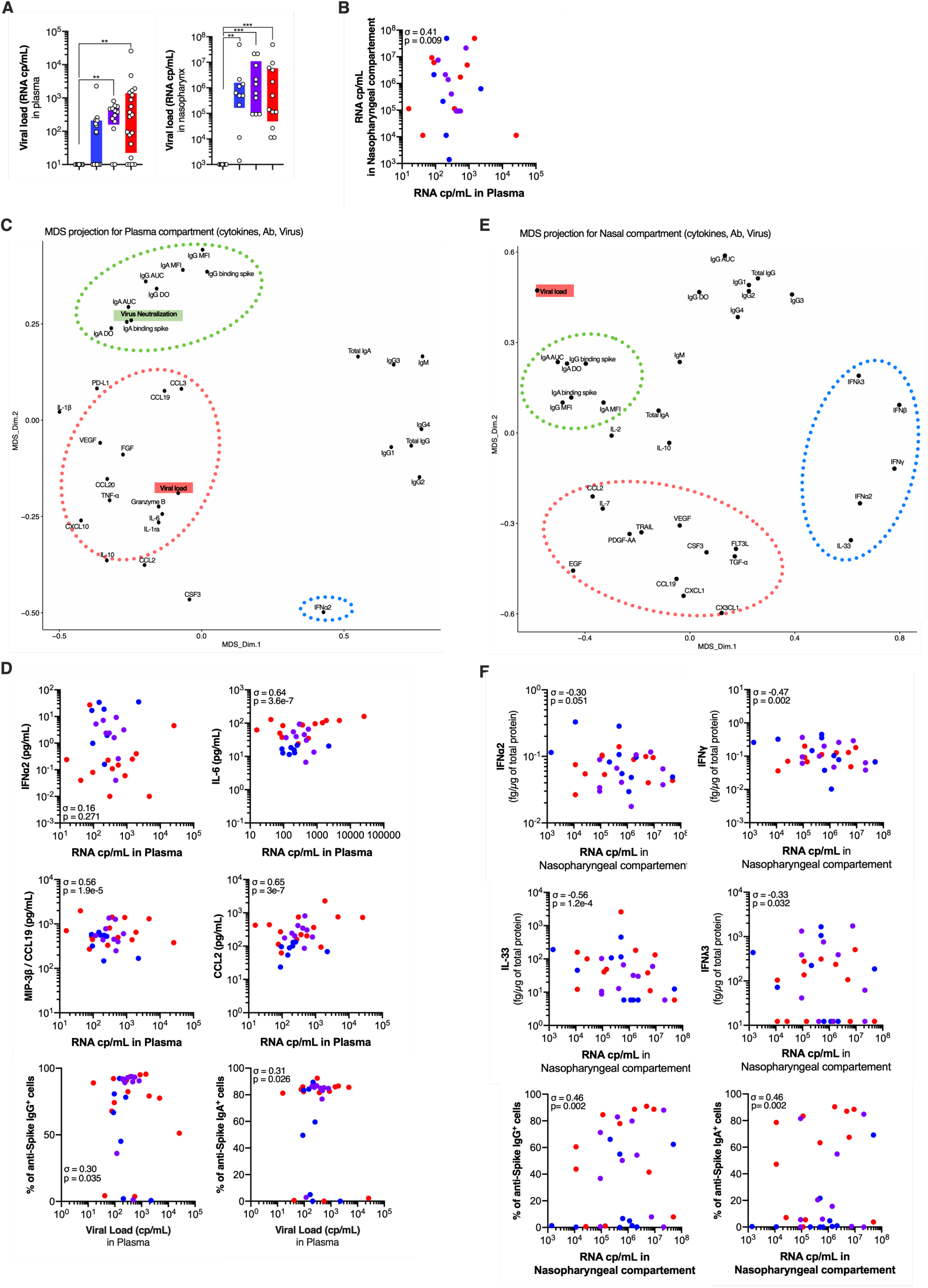
Correlation between cytokines, antibodies and viral load in the plasma and in the mucosa. (**A**) Plasma viral loads evaluated by digital PCR and in nasal swabs estimated by RT-PCR and expressed per cycle threshold (Ct). (**B**) Correlation plots between viral load the in the plasma versus nasopharyngeal compartment. Multidimensional scaling (MDS) projection for (**C**) plasma compartment (cytokines, antibodies, blood viral load) and (**D**) nasopharyngeal compartment (cytokines, antibodies, nasal viral load). (**E**) and (**F**) show individual correlation plots between viral load and cytokines or antibodies. In (**A**), box plots with median ± minimum to maximum. P values were determined with the Kruskal-Wallis test followed by with Dunn’s post-test for multiple group comparisons. In (**B, E** and **F**), σ represents Spearman coefficient and p the p value. *P < 0.05; **P < 0.01; ***P < 0.001.

A similar MDS projection derived from the nasopharyngeal dataset generated a markedly different pattern. In the nasopharynx, SARS-CoV-2 viral loads were more closely associated with spike-specific IgG and IgA responses. However, in contrast with the plasma, viral loads were not positively associated with any inflammatory or regulatory cytokines and showed strong negative correlations with IL-33, IFNγ, IFNλ3, IFNβ and IFNα2 (**Fig. 4E,F; Supplemental Fig. 4B**). These cytokines were decreased in COVID-19 patients (**Fig. 3C, D**) suggesting that their loss could be linked to SARS-CoV-2 infection. In addition, disease severity-associated cytokines (EGF, VEGF, FLT3-L, CXCL1/GROα, PDGF-AA, TGFα) clustered away from other variables indicating distinct regulatory mechanisms (**Fig. 4E; Supplemental Fig. 4B**).

### Microbiome regulation of systemic and mucosal immune responses in SARS-CoV-2

The upper respiratory tract harbors diverse microbial commensal communities that are implicated in protection against disease-causing pathogens (Brugger et al., 2016; de Steenhuijsen Piters et al., 2019). We hypothesized that perturbations in nasopharyngeal microbial profiles might contribute to the diverse outcomes of immune responses and clinical presentation during SARS-CoV-2 infection. We performed unbiased 16S bacterial rRNA sequencing in order to better characterize the commensal communities and potential pathobiont carriage in the nasopharynx of controls and COVID-19 patients (n = 42). V3-V4 region amplicons were sequenced allowing for identification of 464 Operational Taxonomic Units (OTU). Genus level analysis demonstrated significant (p<0.05) perturbations comparing healthy controls to COVID-19 patients (**Fig. 5A**). In addition, analysis of α-diversity (Simpson and Shannon diversity indices; combined measure of evenness and number of bacteria) showed a decrease in 16S rRNA sequences in severe and critical COVID-19 patients (**Fig. 5B**). Richness of microbiota communities (β-diversity) clearly decreased with disease severity and analysis based on Bray-Curtis distance matrix and subjected to principal coordinate analysis suggested that 16S rRNA profiles in critical patients were different from other patients (**Fig. 5C**). A PERMANOVA test showed that nasopharynx microbiota of critical patients is significantly different from healthy controls (**Supplemental Fig. 5A**). Smoking and sex did not affect this clustering (**Supplemental Fig. 5B, C**). We also applied a non-metric multidimensional scaling (NMDS) using Bray-Curtis distances and Partial least squares-discriminant analysis which also showed the critical patients to be different from other patients (**Supplemental Fig. 5D, E**). While nasopharyngeal bacterial load did not change (**Supplemental Fig. 5F**), specific genera showed striking differences (p<0.05) between patients and healthy controls (**Fig 5D,E)** including *Corynebacterium* and *Dolosigranulum* that are thought to provide protection against pathogen and pathobiont invasion (‘beneficial’ commensals; (Brugger et al., 2016; de Steenhuijsen Piters et al., 2019). These were markedly reduced in COVID-19 patients in a severity-dependent fashion (**Fig. 5D,F**). In contrast, the *Staphylococcus* genus and several strict anaerobes (including *Peptostreptococcus* and *Prevotella* genera) were increased in critical COVID-19 patients (**Fig. 5E,G**). These results demonstrate that SARS-CoV-2 infection is associated with perturbations in nasopharyngeal bacterial communities and with accompanying dysbiosis in critical COVID-19 patients.

**Fig. 5.**
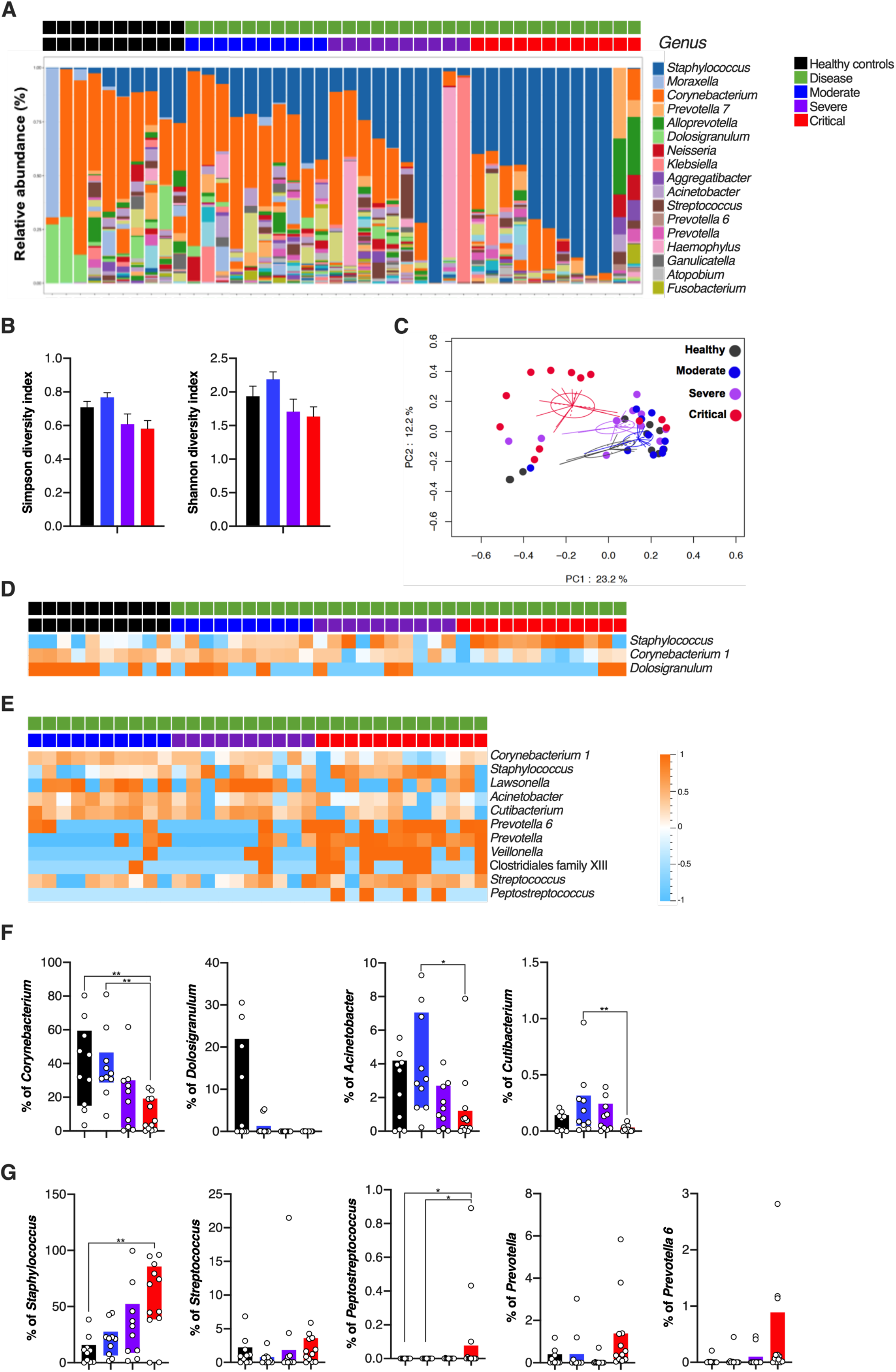
Microbiome 16S evaluation in COVID-19 patients. Nasopharyngeal 16S bacterial communities were measured in the nasopharyngeal compartment of healthy controls (n = 10 donors), mild to moderate (n = 10 patients), severe (n = 10 patients) and critical (n = 12 patients). (**A**) The relative abundance (%) is represented for each *Genus* for each sample. (**B**) Shannon and Simpson diversity indices by patient severity. (**C**) PCA analysis of 16S bacterial profiles (**D**) Heatmap representation of statistically different (P<0.05) *Genus* abundance between healthy controls and COVID-19 patients (moderate, severe, critical). (**E**) Heatmap representation of statistically different (P<0.05) *Genus* abundance between COVID-19 patients depending on disease severity. P values were determined with the Mann-Whitney test. (**F**) and (**G**) Individual *Genus* abundance (%) plots by disease severity. In (**F**) and (**G**), box plots with median ± minimum to maximum. P values were determined with the Kruskal-Wallis test followed by with Dunn’s post-test for multiple group comparisons; *P < 0.05; **P < 0.01; ***P < 0.001.

Finally, we integrated the 16S rRNA bacterial nasopharyngeal microbiome profiles with the immune response (spike-specific antibodies and cytokines) and performed MDS projections in an attempt to undercover associations that might explain mechanistic relationships at this mucosal site. Interestingly, cytokines that decreased with SARS-CoV-2 infection (IL-33, IFNα2 and IFNγ) were associated with the presence of *Dolosigranulum* genus in nasal microbial communities (**Fig. 6A, B; Supplemental Fig. 6A**), whereas IFNλ3 was linked to overall microbial diversity (Shannon, Simpson diversity indices) suggesting genus-specific as well as community-driven regulation of mucosal cytokine production. The previously identified association of viral load and spike-specific antibody responses remained, whereas associations of potential nasal pathobionts (including *Prevotella, Streptococcus, Peptostreptococcus* and *Clostridial* genera) with disease severity-associated nasopharyngeal cytokines was revealed (**Fig. 6A, B; Supplemental Fig. 6A**). MDS projections after integrating nasal microbiome profiles into the plasma datasets revealed intriguing associations of *Staphylococcus* and *Peptostreptococcus* genera with viral load, spike-specific responses, neutralization capacity and inflammatory cytokines (IL-6, TNFα, MIP-3β) (**Fig. 6C, D; Supplemental Fig. 6B**). Taken together, these results reveal an unexpected relationship between nasal microbial communities and local, as well as systemic, cytokine and antibody responses during SARS-CoV-2 infection.

**Fig. 6.**
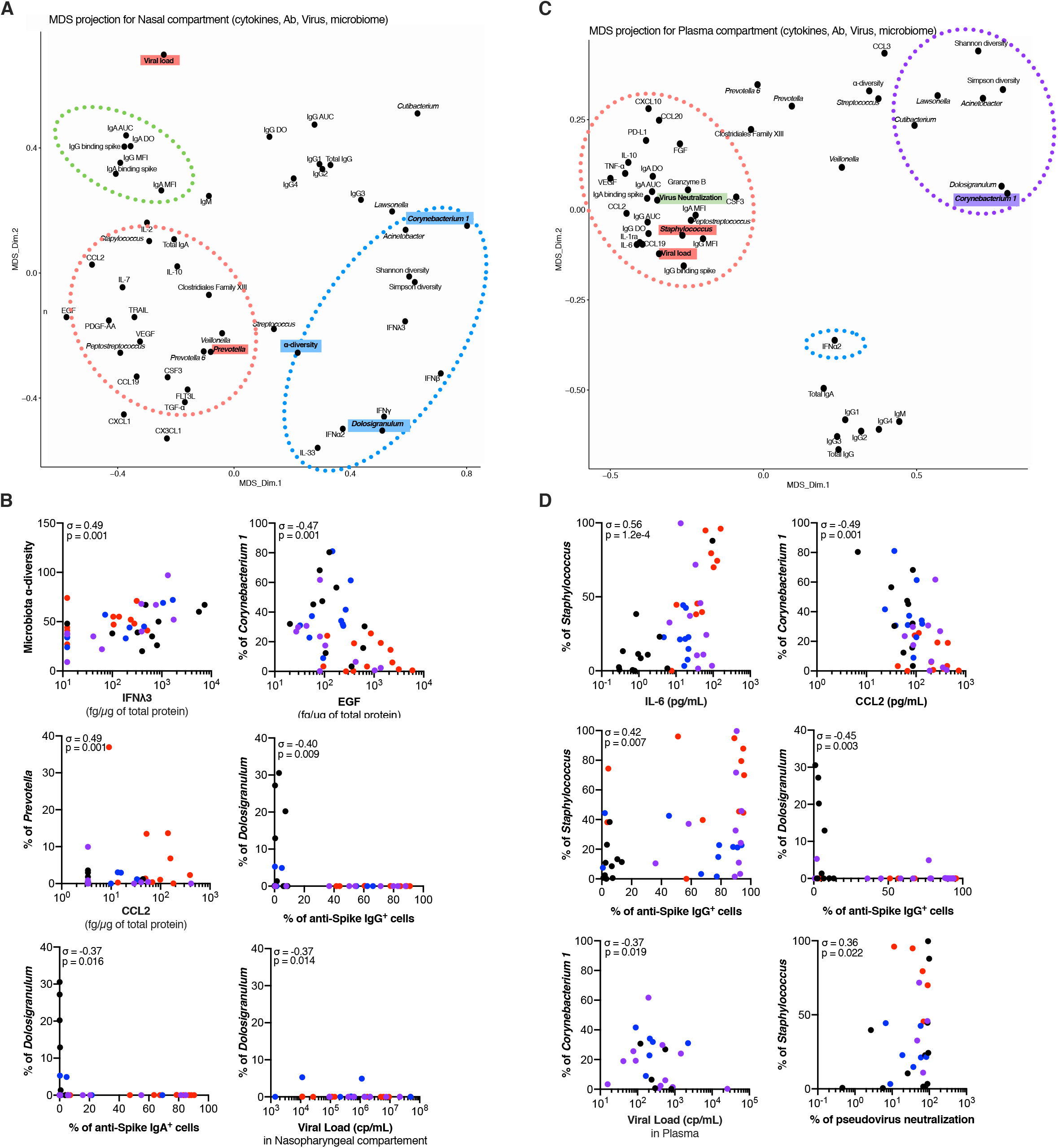
Microbiome regulates mucosal cytokines and antibody responses. Multidimensional scaling (MDS) projection for (**A**) plasma compartment (cytokines, antibodies, blood viral load and nasal microbiome) and (**C**) nasopharyngeal compartment (cytokines, antibodies, nasal viral load and microbiome). (**B**) and (**D**) show individual correlation plots between *Genus* abundance (%) and cytokines, antibodies or viral load. In (**B**) and (**D**), σ represents Spearman coefficient and p the p value.

## Discussion

Despite widespread studies, we still lack a full understanding of how local and systemic immune responses are dysregulated following SARS-CoV-2 infection and the individual roles that they play in determining severe clinical outcomes in a minority of COVID-19 patients. To address this question, we compared systemic and local immune responses during active SARS-CoV-2 infection in a well characterized COVID-19 cohort. We measured host antibody and cytokine responses, determined viral load and characterized nasopharyngeal microbiome using an integrative approach. Comparative analysis of the systemic and local tissue responses suggests a model for protective immunity following SARS-CoV-2 infection and identifies potential regulatory nodes where perturbations may lead to more severe COVID-19 clinical manifestations. First, a healthy nasopharyngeal microbiome (harboring for example ‘beneficial’ components that confer colonization resistance) appears linked to production of nasal cytokines including IL-33, IFNγ, IFNα/β and IFNλ3. SARS-CoV-2 infection, either directly or indirectly, appears to disrupt local microbial homeostasis, resulting in reduced levels of these cytokines that may be important for viral control. Second, while viral load impacts specific humoral immune responses, the local cytokine milieu is also important as evidenced by weaker nasopharyngeal antibody responses in individuals that have lower levels of mucosal inflammatory cytokines. Third, a relative increase in certain bacterial genera associate with enhanced mucosal and systemic inflammation, mediated through distinct cytokine profiles, and correlate with worsening clinical outcome. Together, these findings raise several key questions regarding host mechanisms that can enhance resistance to SARS-CoV-2 infection and associated clinical manifestations.

Resistance to infection by bacterial, fungal and/or viral pathogens is in part mediated through commensal microbial communities that inhabit mucosal surfaces. Several ‘cornerstone’ members contribute to this effect that include *Corynebacterium, Dolosigranulum, Cutibacterium, Lactobacillus and* other genera that generate a ‘front line’ defense against *de novo* infection and suppress progression of ‘pathobionts’ that are present as carriage in normal healthy individuals (Brugger et al., 2016; Claesen et al., 2020; Ridaura et al., 2018; de Steenhuijsen Piters et al., 2019). The mechanisms for this microbial resistance vary and include stimulation of mucus layers and elaboration of antimicrobial peptides (Brugger et al., 2016; Claesen et al., 2020; Ridaura et al., 2018; de Steenhuijsen Piters et al., 2019). How commensal communities participate in anti-viral defense are poorly defined but our results suggest that they may be involved in maintenance of basal production of interferon type I, II and III. Previous studies suggest that microbiota control the constitutive production of type I and type III interferons (Antunes et al., 2019; Di Domizio et al., 2020; Grau et al., 2020; Lan et al., 2019; Schaupp et al., 2020) and modulate the resistance to virus infections in mice (Abt et al., 2012; Bradley et al., 2019; Stefan et al., 2020). Individual variation in microbiome-dependent interferon levels may in part provide an explanation as to differential outcome (resistance versus productive infection and potential spread) after SARS-CoV-2 encounter.

Analysis of the nasopharyngeal antibody response also revealed highly heterogenous responses. While the vast majority of patients generated systemic viral specific antibodies, a surprisingly high proportion of patients had neither detectable viral specific IgG or IgA in their nasopharyngeal compartments despite the use of highly sensitive assays. The presence of nasopharyngeal antibodies was associated with local viral load but also appeared to be regulated by local inflammation and cytokine production. As our study relied on a single sample (taken at day 8-12 post symptom onset), we cannot exclude later ‘nasoconversion’ in patients lacking mucosal spike-specific antibodies. Additional studies involving replication cohorts and longitudinal sampling may be required to determine mechanistic interactions. Recently IgA, and in particular dimeric IgA, were shown to have the most potent neutralizing activity against SARS-CoV-2 (Wang et al., 2020) especially in the early phase of infection (Sterlin et al., 2020). Understanding mechanisms that allow for efficient up-regulation of local IgA production (as in type ‘C’ individuals) and local viral control (Weitnauer et al., 2016) may provide new avenues for protection against SARS-CoV-2. In light of these and our own findings, nasopharyngeal immune response should be considered as potential biomarkers for correlates of protection during SARS-CoV-2 vaccination campaigns.

Multiple studies have recently described a parallel impaired type I interferon activity and exacerbated inflammatory cytokine response in severe COVID-19 (Blanco-Melo et al., 2020; Hadjadj et al., 2020). While systemic hyperinflammation is likely detrimental for an uneventful clinical recovery, such responses may be required during initial infection as reflected by the poor outcome of clinical studies targeting these cytokine pathways (anti-IL-6, IL-1 therapies) during the early phase of disease (Huang et al., 2020a). Furthermore, while the importance of type I interferons has been demonstrated through multiple lines of evidence, including both genetic variants and presence of neutralizing antibodies in severe patients (Bastard et al., 2020; Zhang et al., 2020), some uncertainty remains in the literature possibly due to differences in the site of analysis, methods used, or the time point studied (Galani et al., 2020; Lee and Shin, 2020; Lee et al., 2020; Lucas et al., 2020) as well as their lack of efficacy in randomized placebo control trials (WHO Solidarity Trial Consortium et al., 2020). As such, a comparative in-depth analysis of local and systemic inflammatory cytokines appeared warranted in order to uncover potential mechanisms that might regulate disease severity in COVID-19 patients. The main overlap between mucosal and systemic compartments was an increase in CCL2, a critical cytokine for recruitment of monocytes to infected and inflamed tissues. These findings support previous reports indicating a general critical role for this cytokine in COVID-19 (Lucas et al., 2020; Sierra et al., 2020) which was elevated in bronchoalveolar lavage fluid from the lungs of Covid-19 patients during mechanical ventilation (Zhou et al., 2020b). Furthermore, a recent GWAS analysis identified a polymorphism in the CCL2 receptor (CCR2) as associated with critical COVID-19 illness (Pairo-Castineira et al., 2020).

However, the main striking result from our study was significant elevations in critical patients of a cluster of plasma cytokines and growth factors that did not have an obvious role in anti-viral immunity. Insight into their implication in severe COVID-19 came from the integration of plasma datasets with the nasal microbiome which revealed positive associations with opportunistic bacterial genera such as *Prevotella* and *Streptococcus*, and negative associations with key mucosal cornerstone genera such as *Corynebacterium* and *Dolosigranulum*. This hypothesis was supported by additional associations between presence of nasopharyngeal *Staphylococcus* genus and plasma inflammatory cytokines such as IL-6. Whether such nasal dysbiosis drives systemic inflammation will require further study, but this hypothesis has further support from a recent study documenting a similar mechanism in the infected intestine (Giron et al., 2020). Previous studies have document ‘pathobiont’ carriage (including *Staphylococcus aureus, Streptococcus pneumoniae* and *Haemophilus influenzae*) in up to 40% of healthy individuals (Brugger et al., 2016). Our results suggest that these individuals may be at higher risk of developing severe COVID-19 disease as SARS-CoV-2 infection would result in a breakdown of local epithelial barrier function leading to escape of these potential pathobionts with resultant systemic manifestations. In summary our study identifies novel host-viral-microbiome interactions during infection with SARS-CoV-2 which may help new strategies for identifying at risk individuals.

## Data Availability

All data are available in the main text or the supplementary materials.

## Acknowledgments

This study was supported by an Institut Pasteur Covid-19 research grant and by a grant (CoVarImm) from the Agence National de la Recherche (ANR-flash Covid19) awarded to DD and JPD, and by the Laboratoire d’Excellence ‘‘Milieu Intérieur” (grant no. ANR-10-LABX-69-01) and the Fonds IMMUNOV, for Innovation in Immunopathology. NS is a recipient of the Pasteur-Roux-Cantarini Fellowship. We thank the UTechS CB of the Center for Translational Research, Institut Pasteur for supporting Luminex and Simoa analysis. We thank Laurence Motreff and Laurence Ma, Biomics Platform, C2RT, Institut Pasteur, Paris, France, supported by France Génomique (ANR-10-INBS-09-09), IBISA and the Illumina COVID-19 Projects’ offer for microbial sequencing. We thank Amine Ghozlane, Bioinformatics and Biostatistics HUB, Institut Pasteur, Paris, for the assistance with the 16S rRNA sequencing data analysis. We acknowledge all health-care workers involved in the diagnosis and treatment of patients in Cochin Hospital, especially Célia Azoulay Lauren Beaudeau, Etienne Canoui, Pascal Cohen, Adrien Contejean, Bertrand Dunogué, Didier Journois, Paul Legendre, Jonathan Marey and Alexis Régent.

## Author Contributors

NS, PG, DD and JPD conceived and designed the study, wrote the manuscript and had full access to all of the data in the study and take responsibility for the integrity of the data and the accuracy of the data analysis. NS, PG, BC, VR, DD, JPD, OS, TB, and HM performed data analysis. MB, CP, TB, LG, VB, HP, SHM performed specific experimental analysis. NY, JH, SK, FRL, and BT were involved in the clinical study and sample collection. All authors critically revised the manuscript for important intellectual content and gave final approval for the version to be published. All authors agree to be accountable for all aspects of the work in ensuring that questions related to the accuracy or integrity of any part of the work are appropriately investigated and resolved.

## Declaration of interests

We declare no competing interests.

## STAR Methods

### Study design

This non-interventional study was conducted between March 19, 2020 and April 3, 2020 in Cochin Hospital (Paris, France), in the setting of the local RADIPEM biological samples collection, derived from samples collected in routine care as previously described (Hadjadj et al., 2020). Biological collection and informed consent were approved by the Direction de la Recherche Clinique et Innovation (DRCI) and the French Ministry of Research (N°2019-3677). Inclusion criteria for COVID-19 inpatients were: age between 18 and 80 years old, diagnosis of COVID-19 according to WHO interim guidance, and positive SARS-CoV-2 RT-PCR testing on a respiratory sample (nasopharyngeal swab or invasive respiratory sample). Inpatients with pre-existing unstable chronic disorders (such as uncontrolled diabetes mellitus, severe obesity defined as body mass index greater than 30, unstable chronic respiratory disease or chronic heart disease) and with bacterial co-infection were excluded. Since median duration from onset of symptoms to respiratory failure was previously shown to be 9.5 (interquartile range, 7.0-12.5) days (Yang et al., 2020), we analyzed immune responses between 8 to 12 days after onset of first symptoms for all patients and before the initiation of any antiviral or anti-inflammatory treatment. Healthy controls were asymptomatic adults, matched with cases on age (+/-5 years), with a negative SARS-CoV-2 RT-PCR testing at time of inclusion. The study conforms to the principles outlined in the Declaration of Helsinki, and received approval by the appropriate Institutional Review Board (Cochin-Port Royal Hospital, Paris, France; number AAA-2020-08018).

Epidemiological, demographic, clinical, laboratory, treatment, and outcome data were extracted from electronic medical records using a standardized data collection form. Chest radiographs or CT scan were also done for all inpatients. Laboratory confirmation of SARS-CoV-2 was performed at the Cochin Hospital, Virology Department, Paris, France. RT-PCR assays were performed in accordance with the protocol established by the WHO (World Health Organization. Coronavirus disease (COVID-19) technical guidance: laboratory testing for 2019-nCoV in humans (https://www-who-int.proxy.insermbiblio.inist.fr/emergencies/diseases/novel-coronavirus-2019/technical-guidance/laboratory-guidance).

The severity of COVID-19 was classified at the time of admission based on the adaptation of the Sixth Revised Trial Version of the Novel Coronavirus Pneumonia Diagnosis and Treatment Guidance. Mild cases were defined as mild clinical symptoms (fever, myalgia, fatigue, diarrhea) and no sign of pneumonia on thoracic computed tomography (CT) scan. Moderate cases were defined as clinical symptoms associated with dyspnea and radiological findings of pneumonia on thoracic CT scan, and requiring a maximum of 3 L/min of oxygen, stable for at least the following 24 hours. Severe cases were defined as respiratory distress requiring more than 3 L/min of oxygen and no other organ failure, stable for at least the following 24 hours. Critical cases were defined as respiratory failure requiring mechanical ventilation, shock and/or other organ failure that require an intensive care unit (ICU).

### Patient characteristics

Forty-nine Covid-19 patients and 12 healthy controls were included. The demographic and clinical characteristics of the patients have been previously described (Hadjadj et al., 2020). The median age of the patients was 55 years (interquartile range, 50 to 63) and 78% were male, while median age of healthy controls was 51 years (interquartile range, 38 to 60) and 72% were male. Patients were sampled for plasma and nasal swabs after a median duration of 10 days (interquartile range, 9 to 11) after disease onset. Fever was present in 98% of the patients, and other most common symptoms were dyspnea (98%), fatigue (96%), cough (92%), myalgia (62%) and diarrhea (34%). Among the whole population, 44% had at least one controlled coexisting illness, mainly hypertension and type 2 diabetes.

On admission, the degree of severity of Covid-19 was categorized as moderate in 15 patients (median oxygen requirement 2 L/min), severe in 11 patients (median oxygen requirement 5 L/min) and critical in 23 patients. Of CT scan available at the time of admission, all were abnormal, showing ground-glass opacities (100%) with bilateral patchy distribution (96%). Most of the patients had elevated CRP, ferritin and lactate dehydrogenase (LDH) levels. Patients with severe and critical disease had more prominent laboratory abnormalities than those with mild-to-moderate disease, and extension on chest CT scan was correlated with disease severity. No patients with moderate disease required admission to an ICU or the use of mechanical ventilation, while 6 out of 11 patients with severe disease were eventually admitted to the ICU.

### Nasopharynx swab processing

Nasopharynx swabs were thawed in a P3 laboratory and vortexed for 1 minute at 2500 rpm to ensure complete sample recovery. Samples (1 mL media) were transferred in a 96-well deep-well plate and centrifuged at 16,000 g for 10 minutes at 4°C to pellet the cells and accompanying microbes for 16S rRNA analysis. Supernatants were recovered and either heat-inactivated for antibody analysis, or S/D treated for cytokine analysis as described below. Total protein determinations were performed using the Bio-Rad Protein Assay (Bradford, 1976) with serum albumin as standard.

### Antibody assays

SARS-CoV-2 specific antibodies were quantified using assays previously described (Grzelak et al., 2020). Briefly, a standard ELISA assay using as target antigens the extracellular domain of the S protein in the form of a trimer (ELISA tri-S) and the S-Flow assay, which is based on the recognition of SARS-CoV-2 S protein expressed on the surface of 293T cells (293T-S), were used to quantify SARS-CoV-2 specific IgG and IgA subtypes in plasma and nasal swab supernatants. Assay characteristics including sensitivity and specificity were previously described (Grzelak et al., 2020). Total IgA, IgM, IgG1, IgG2, IgG3 and IgG4 were determined using the Bio-Plex Pro Human Isotyping Assay Panel (Biorad, Hercule, CA, USA) according to the manufacturers’ instructions.

### Cytokine assays

Prior to protein analysis plasma and nasal samples were treated in a P3 laboratory for viral decontamination using a protocol previously described for SARS-CoV (Darnell and Taylor, 2006) which we validated for SARS-CoV-2. Briefly, samples were treated with TRITON X100 (TX100) 1% (v/v) and Tri-n-Butyl Phosphate (TnBP) 0.3% (v/v) for 2hrs at RT. TnBP was removed prior to cytokine analysis by passing the treated samples though C18 columns. IFNα2, IFNγ, IL-17A, (triplex) and IFNβ and IFNλ3 (both single plex) protein plasma and nasal concentrations were quantified by Simoa assays developed with Quanterix Homebrew kits as previously described (Rodero et al., 2017). IL-6, TNFα, and IL-10 were measured with a commercial triplex assay (Quanterix). For the IFNα2 assay, the BMS216C (eBioscience) antibody clone was used as a capture antibody after coating on paramagnetic beads (0.3mg/mL), and the BMS216BK already biotinylated antibody clone was used as the detector at a concentration of 0.3ug/mL. The SBG revelation enzyme concentration was 150pM. Recombinant IFNα2c (eBioscience) was used as calibrator. For the IFNγ assay, the MD-1 antibody clone (BioLegend) was used as a capture antibody after coating on paramagnetic beads (0.3 mg/mL), the 25718 antibody clone (R&D Systems) was biotinylated (biotin/antibody ratio = 40/1) and used as the detector antibody at a concentration of 0.3ug/mL. The SBG revelation enzyme concentration was 150pM. Recombinant protein (PBL Assay Science) was used to quantify IFNγ concentrations. For the IL-17A assay, the BL23 antibody clone (BioLegend) was used as a capture antibody after coating on paramagnetic beads (0.3 mg/mL), the MT504 antibody clone (MabTech), already biotinylated, was used as the detector antibody at a concentration of 0.3ug/mL. The SBG revelation enzyme concentration was 150pM. For the IFNβ assay, the 710322-9 IgG1, kappa, mouse monoclonal antibody (PBL Assay Science) was used as a capture antibody after coating paramagnetic beads (0.3 mg/mL), the 710323-9 IgG1, kappa, mouse monoclonal antibody (PBL Assay Science) was biotinylated (biotin/antibody ratio = 40/1) and used as the detector antibody, and recombinant protein (PBL Assay Science) were used to quantify IFNβ concentrations. For the IFNλ3 assay, the MMHL-3 IgG1 kappa mouse monoclonal antibody (PBL Assay Science) was used as a capture antibody after coating paramagnetic beads (0.3 mg/mL), the 567107R IgG2a mouse monoclonal antibody (R&D systems) was biotinylated (biotin/antibody ratio = 60/1) and used as the detector antibody, and recombinant protein (PBL Assay Science) were used to quantify IFNλ3 concentrations. The limit of detection (LOD) of these assays were 0.6 pg/mL for IFNβ, 0.6 pg/mL for IFNλ3, 2 fg/mL for IFNα, 7 fg/mL for IFNγ and 3 pg/mL for IL17A including the dilution factor. An additional 38 cytokines and chemokines were measured in plasma and nasal supernatants with a commercial Luminex multi-analyte assay (Biotechne, R&D systems).

### Quantification of Nasopharyngeal viral load

Nasopharyngeal viral loads were determined using RdRp-IP4 quantitative RT-PCR designed at the Institut Pasteur (National Reference Center for Respiratory Viruses) to target a section of the RdRp gene based on the first sequences of SARS-CoV-2 made available on the Global Initiative on Sharing All Influenza Data database on Jan 11, 2020 (Lescure et al., 2020). Primer and probe sequences: nCoV_IP4-14059Fw GGTAACTGGTATGATTTCG; nCoV_IP4-14146Rv CTGGTCAAGGTTAATATAGG; nCoV_IP4-14084Probe(+) TCATACAAACCACGCCAGG [5’]Fam [3’]BHQ-1. All positive samples were quantified using a standard curve and expressed as number of RNA copies per mL.

### Quantification of plasma viral load

SARS-CoV-2 viremia was quantified in each patient blood sample using the Naica^™^ droplet based digital PCR (ddPCR) machine Stilla^Inc^ (Villejuif, France) with COVID-19 Multiplex Crystal digital PCR detection kit (Apexbio^Inc^, Beijing, China) as previously described (Veyer et al., 2020). Plasma viral RNA was extracted using QIAamp® Viral RNA Mini Kit following the manufacturer guidelines. Results were automatically analyzed using “Crystal reader” and “Crystal Miner” software and SARS CoV-2 viral concentration (cp/mL) were finally calculated considering the extracted volume of plasma (140 µL).

### S-Pseudotype neutralisation assay

293T cells stably expressing ACE2 (293T-ACE2) were made by lentiviral transduction and selection with puromycin (1 ug/mL). To perform the assay, 2×10^4^ cells were detached with PBS-EDTA and seeded in Flat-bottom 96-well plates. S-pseudotypes were incubated with the sera to be tested (at 1:100 dilution, unless otherwise specified) in culture medium, incubated 15 min at RT and added on transduced cells. After 48 hours Bright-Glo ™ Luciferase Assay System was added to the wells and the luciferase signal was measured with EnSpire^®^ Multimode Plate Reader (PerkinElmer). The percentage of neutralization was calculated as follow: 100 x (1 − mean (luciferase signal in sample duplicate)/mean(luciferase signal in virus alone)). S-pseudotypes incubated without serum and medium alone were used as positive and negative controls, respectively.

### Bacterial DNA isolation and 16S rRNA sequencing

We extract total genomic DNA from swabs samples, using the NucleoSpin® 96 Genomic DNA kit (Macherey-Nagel ®). Negative control samples included buffers only. Briefly, the pellets were incubated with Ready-Lyse™ Lysozyme Solution (250 U/μl) (Epicentre, Hessisch Oldendorf, Germany) for 30 minutes at 37°C followed Proteinase K digestion buffer at 55°C overnight. Carrier (20 μg glycogen) was added and DNA extraction was performed as per manufacturers’ instructions. DNA was eluted in 25 μl and immediately frozen at −80°C. The concentration of extracted DNA is determined using TECAN (QuantiFluor® ONE dsDNA System, Promega), and DNA integrity and size were also confirmed with the Agilent 2100 Bioanalyzer (Agilent Technologies, USA). The V3-V4 region of bacterial 16S rRNA was amplified using V3-340F (CCTACGGRAGGCAGCAG) and V4-805R (GGACTACHVGGGTWTCTAAT) primers (Chakravorty, Helb et al. 2007, Castelino, Eyre et al. 2015). The primers have a primer linker, primer pad, unique 8-mer Golay barcode which was used to tag PCR products from respective samples and negative control. PCR reactions consisted of 18 μl of AccuPrime Pfx Super Mix (12344-040; Invitrogen), 0.5 μl of each primers and 1 μl of DNA (10 ng). PCR was carried out as follows: 95 °C for 2 min, 30 cycles of 95 °C for 20 s, 55 °C for 15 s and 72 °C for 1 min, and a final extension step at 72 °C for 10 min on a Biorad thermocycler. PCR products were cleaned using Nucleo Mag magnetic purification beads (MACHEREY-NAGEL Kit) following the protocol, quantified with the Quanti Fluor® ONE dsDNA kit (Promega), and pooled in equal amounts of each PCR product. Library pools were loaded at 12pM with a 15% PhiX spike for diversity and sequencing control, onto a v3 300-bp paired end reads cartridge for sequencing on the Illumina MiSeq NGS platform.

### Sequence processing and statistical analysis

After removing reads containing incorrect primer or barcode sequences and sequences with more than one ambiguous base, we recovered from 42 samples a total of 8.075.384 reads (192.271 reads on average). The bioinformatics analysis was performed as previously described (Volant et al., 2020). Briefly, amplicons were clustered into operational taxonomic units (OTU) with VSEARCH (v1.4) and aligned against the SILVA database. The input amplicons were then mapped against the OTU set to get an OTU-abundance table containing the number of reads associated with each OTU. The normalization, statistical analyses and multiple visualization were performed with SHAMAN (SHiny application for Metagenomic Analysis (shaman.c3bi.pasteur.fr) based on R software.

### Bacterial load quantification by qPCR

To gain further insight into microbiota counts, a qPCR was applied, using universal 16S rRNA primers to measure total bacteria (16S_F: 5’-ATTACCGCGGCTGCTGG-3’ and 16S and 16S_R: 5’-ATTACCGCGGCTGCTGG-3’) and 18S rRNA primers to measure total fungi (18S_F: 5’-ATTGGAGGGCAAGTCTGGTG-3’ and 18S_R: 5’-CCGATCCCTAGTCGGCATAG-3’) (Qiu, Zhang et al. 2015). PCR reactions consisting of 10 μL SYBR Green PCR master mix (Roche), 1 μL (10 nM) each primer, 200 ng template cDNA in 20 μL of reaction carried out on an ABI StepOnePlus Sequence Detection System (Applied Biosystems). Thermocycling reactions consisted of 1 min at 95 °C followed by 40 cycles of 15 s at 95 °C, 15 s at 56 °C, and 45 s at 72 °C.

### Statistical analysis

GraphPad Prism was used for statistical analysis. Cytokines were filtered first on variance (σ>2) to remove analytes where the majority of values were at or close to the limit of detection (LOD). P values were determined by a Kruskal-Wallis test, followed by Dunn’s post-test for multiple group comparisons with median reported; *P < 0.05; **P < 0.01; ***P < 0.001. Correlations between the different assays were calculated using Spearman test. Heatmaps were generated with Qlucore OMICS explore Version 3.5(26). Correlation matrices were built using the Spearman correlation, and computed using R (v4.0.3). Correlation plots were generated with R package `corrplot`(v0.84). The Multi-Dimensional Scaling (MDS) plots were derived from the correlation matrices by defining a similarity metric equal to `1-Rs(a,b)` where Rs(a,b) is the spearman correlation between factor a and b. MDS computation was performed with the ‘cmdscale’ function from the `stats`package (v4.0.3). Plots were made using the ggplot2 package (v3.3.2).

## Notes

### Competing Interest Statement

The authors have declared no competing interest.

### Author Declarations

The study was performed in accordance with the declaration of Helsinki, and received approval by the appropriate Institutional Review Board, (Cochin-Port Royal Hospital, Paris, France) approval number AAA-2020-08018.

