## Supplemental Information for "Distinct systemic and mucosal immune responses to SARS-CoV-2"

**Supplementary Fig. 1. Systemic and mucosal antibody responses in COVID-19 patients.** (**A**) Correlation plots between the anti-Spike IgA/IgG OD ELISA and the anti-Spike IgA or IgG responses (% by S_flow) in plasma (**A**) or in nasopharyngeal compartment (**C**). Total IgM, IgG and IgA and IgG1/2/3/4 were measured in plasma (**B**) or in nasopharyngeal compartment (**D**) using a bead-based multiplexed immunoassay system Luminex. In (**A**) and (**C**), σ represents Spearman coefficient and p the p value. In (**B**) and (**D**), box plots with median ± minimum to maximum. P values were determined with the Kruskal-Wallis test followed by with Dunn's post-test for multiple group comparisons; *P < 0.05.

**Supplemental Fig. 2. Compartmentalized spike-specific antibody responses in COVID-19.** (**A**) Correlation plots between the anti-Spike IgA or IgG OD ELISA with the anti-Spike IgA or IgG responses (% by S_flow) in plasma and nasopharynx. (**B**) Correlation plots between the anti-Spike IgA or IgG D.O. ELISA in plasma versus nasopharynx. (**C**) Correlation plots between plasma and nasopharynx anti-Spike IgA/IgG D.O. ELISA or anti-Spike IgA or IgG responses (% by S_flow) in plasma versus nasopharynx. (**D**) Correlation plots between plasma IgA versus nasopharynx IgG responses. (**E**) Heatmap representation of all antibodies measured in plasma and nasopharyngeal compartment. In (**A, B, C**), σ represents Spearman coefficient and p the p value.

**Supplemental Fig. 3. Systemic and mucosal cytokines in COVID-19 patients.**
(**A**) Heatmap representation of statistically different (P<0.05) plasma cytokines between critical COVID-19 patients and mild to moderate and severe COVID-19 patients. (**B**) Plasma cytokine concentration plots by patient severity. (**C**) Heatmap representation of statistically different (P<0.05) nasopharyngeal cytokines between critical COVID-19 patients and mild to moderate and severe COVID-19 patients. (**D**) Nasopharyngeal cytokine concentration plots by patient severity. (In (**A**) and (**C**), P values were determined with the Mann-Whitney test. In (**B**) and

(**D**), box plots with median ± minimum to maximum. P values were determined with the Kruskal-Wallis test followed by with Dunn's post-test for multiple group comparisons; *P < 0.05; **P < 0.01; ***P < 0.001.

**Supplemental Fig. 4. Correlation between cytokines, antibodies and viral load in the plasma and in the mucosa.** Correlation matricies between the (**A**) systemic compartment (plasma cytokines, antibodies, blood viral load) and (**B**) nasopharyngeal compartment (cytokines, antibodies, nasal viral load).

**Supplementary Fig. 5. Microbiome 16S evaluation in COVID-19 patients.** (**A**) PERMANOVA test analysis of 16S bacterial profiles, PCA analysis of 16S bacterial profiles and color coded by smoking status (**B**) and sex (**C**). (**D**) Non-metric multidimensional scaling (MDS) of 16S bacterial profiles. (**E**) Partial least squares-discriminant analysis of 16S bacterial profiles (**F**) Bacterial load (16S rRNA) plotted by patient severity. (**G)** Individual correlation plots between *Genus* abundance (%) for 'cornerstone' and 'pathobionts'.

**Supplemental Fig. 6. Microbiome regulates mucosal cytokines and antibody responses.** Correlation matricies between the (**A**) systemic compartment (cytokines, antibodies, blood viral load and nasal microbiome) and (**B**) nasopharyngeal compartment (cytokines, antibodies, nasal viral load and microbiome).
