## Supplementary figures and images for "Distinct systemic and mucosal immune responses to SARS-CoV-2"

### Supplemental Figure 1

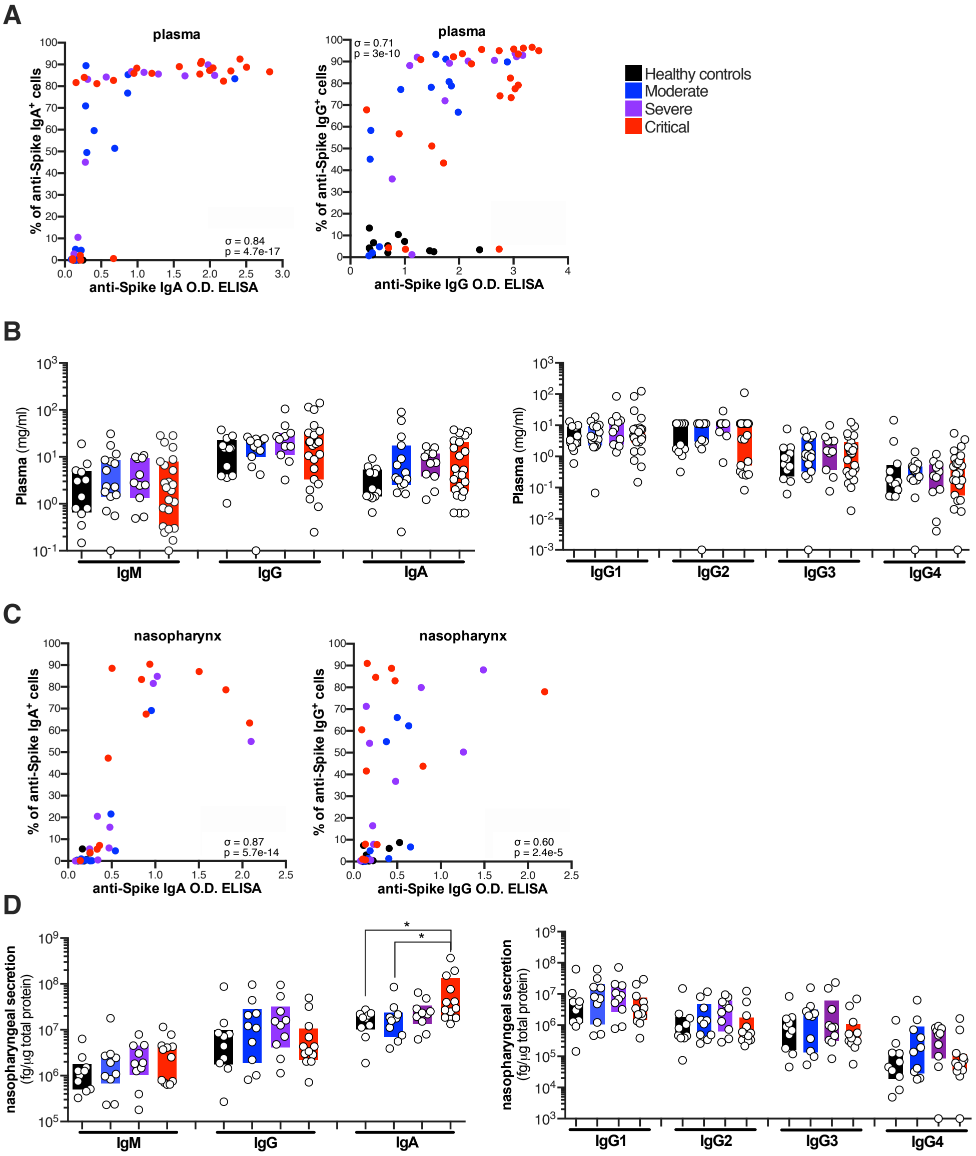

### Supplemental Figure 2

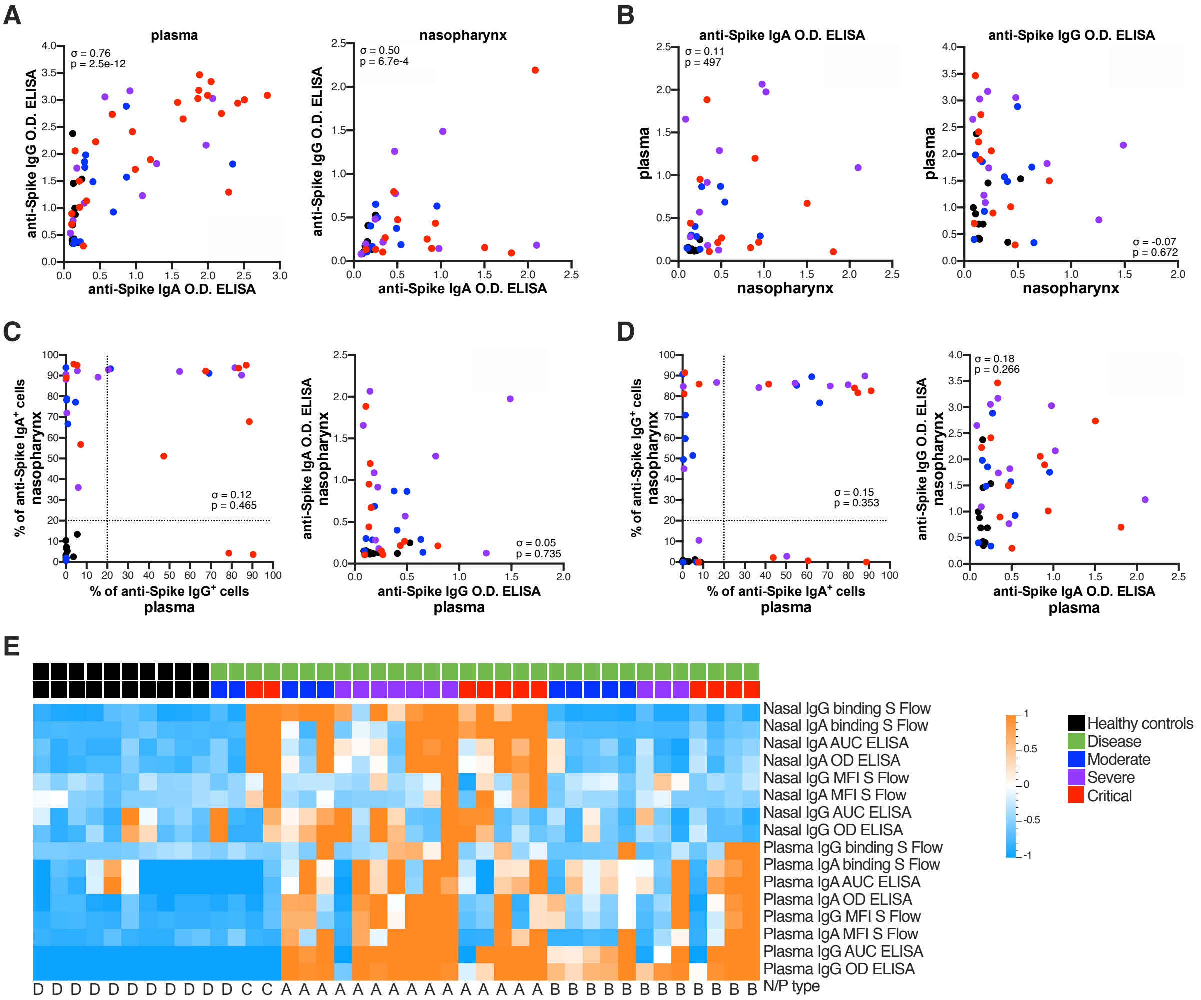

### Supplemental Figure 3

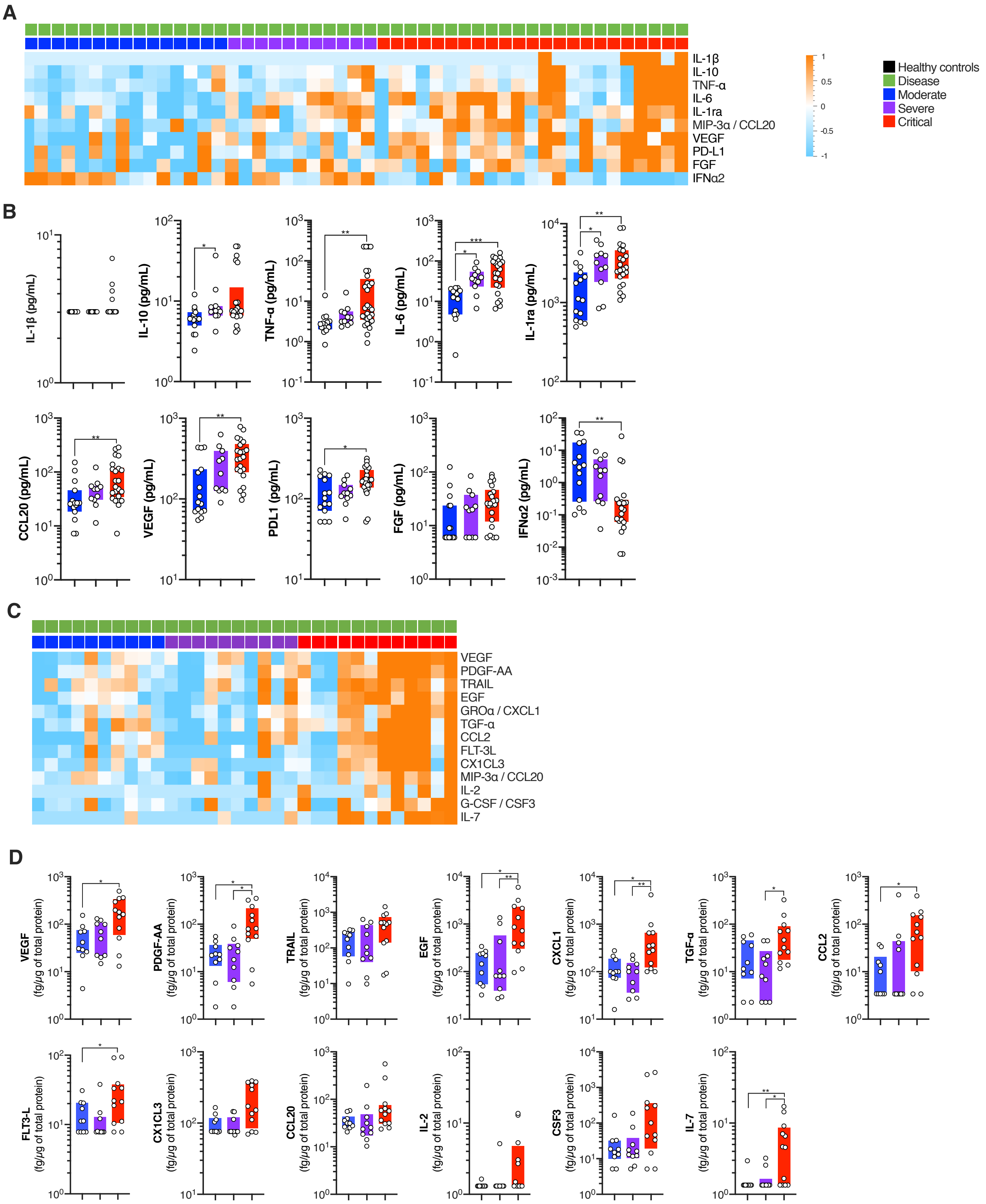

### Supplemental Figure 4

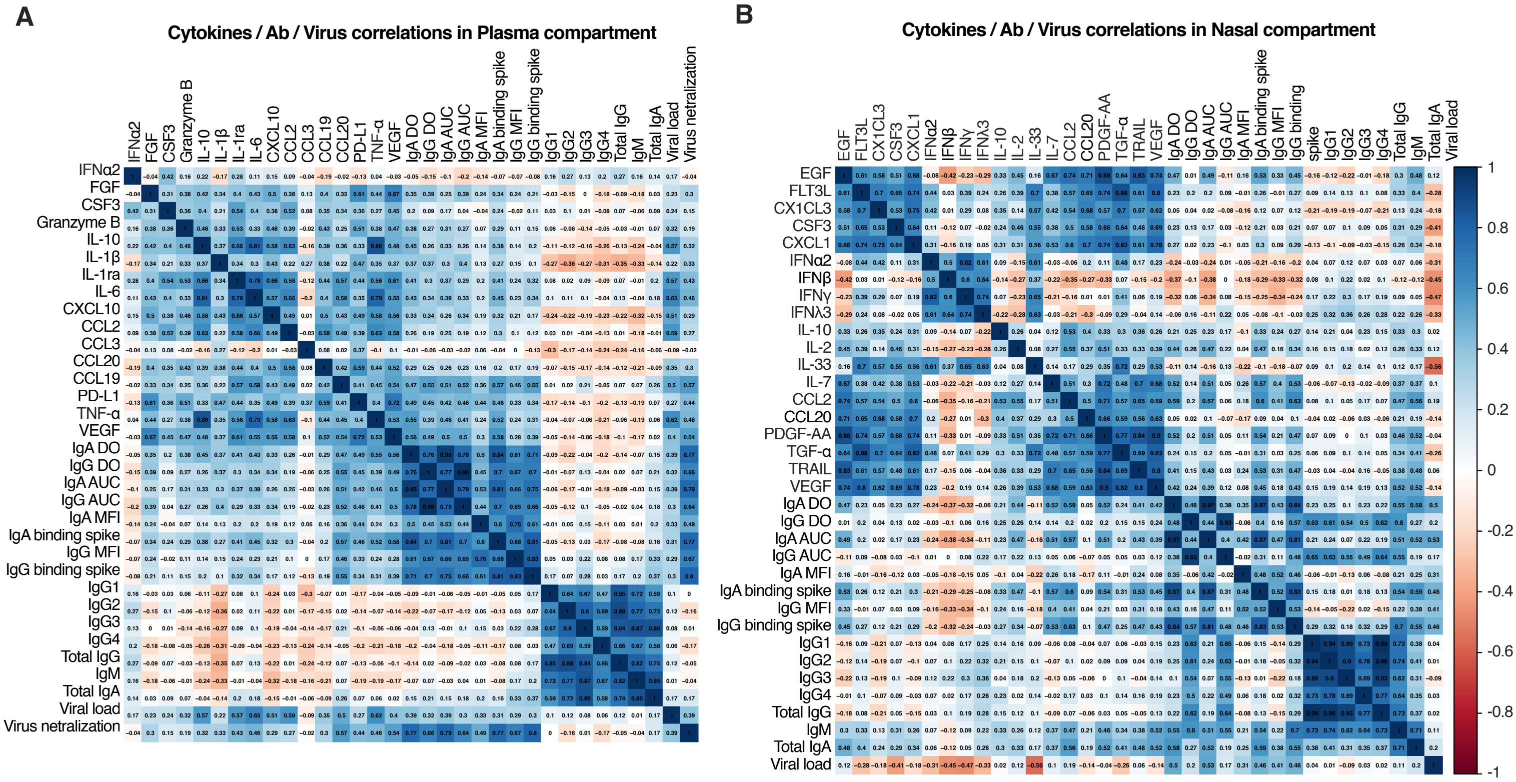

### Supplemental Figure 5

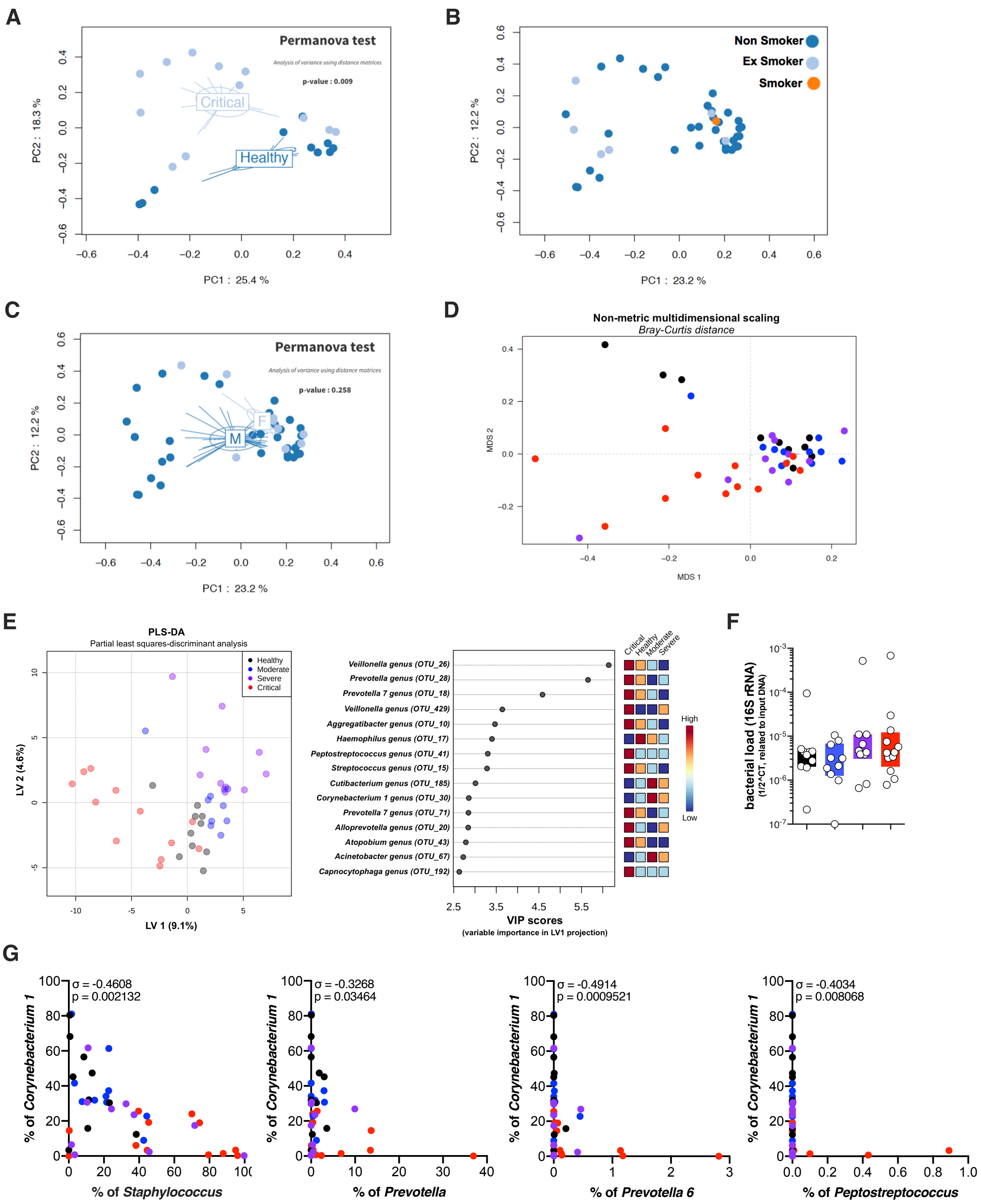
