## Supplemental Figure 6 for "Distinct systemic and mucosal immune responses to SARS-CoV-2"

A

### Cytokines/Ab/Virus in Plasma compartement with Nasal microbiome

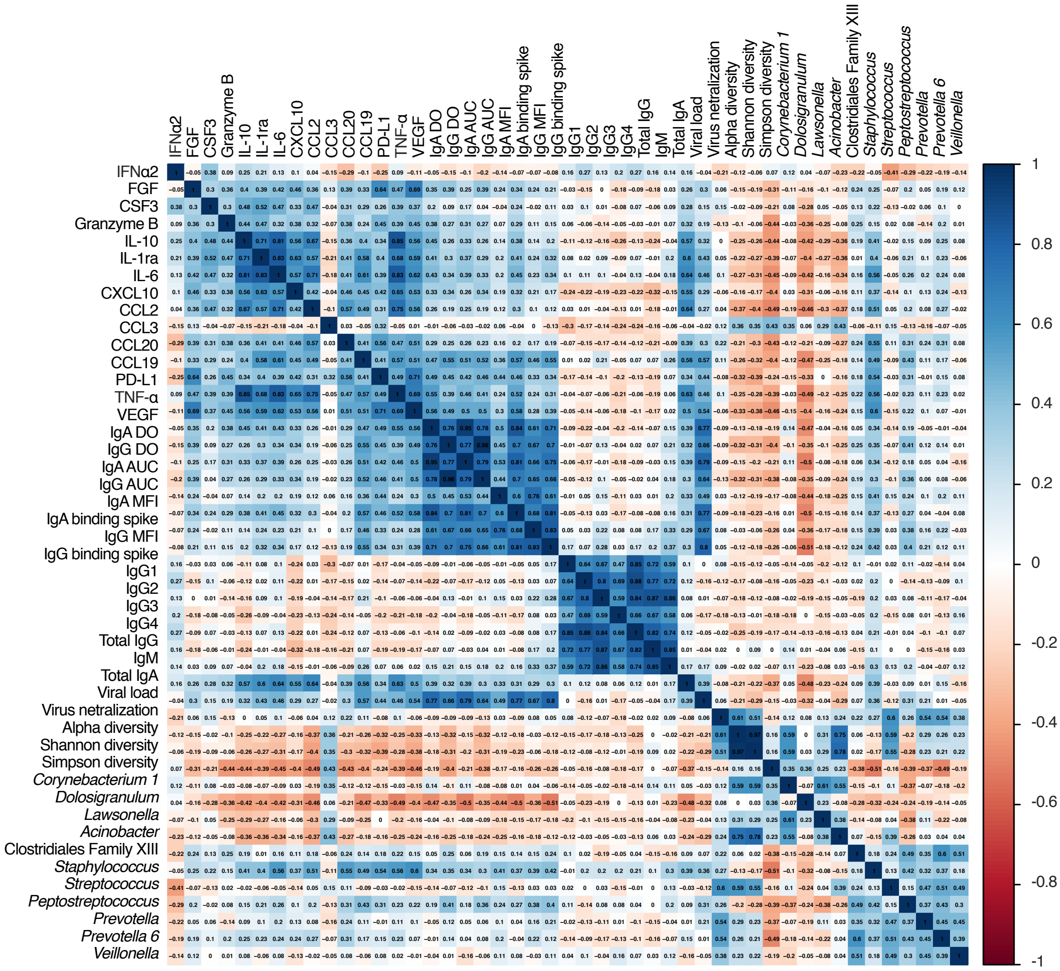


B

### Cytokines/Ab/Virus/Microbiome in Nasal compartement

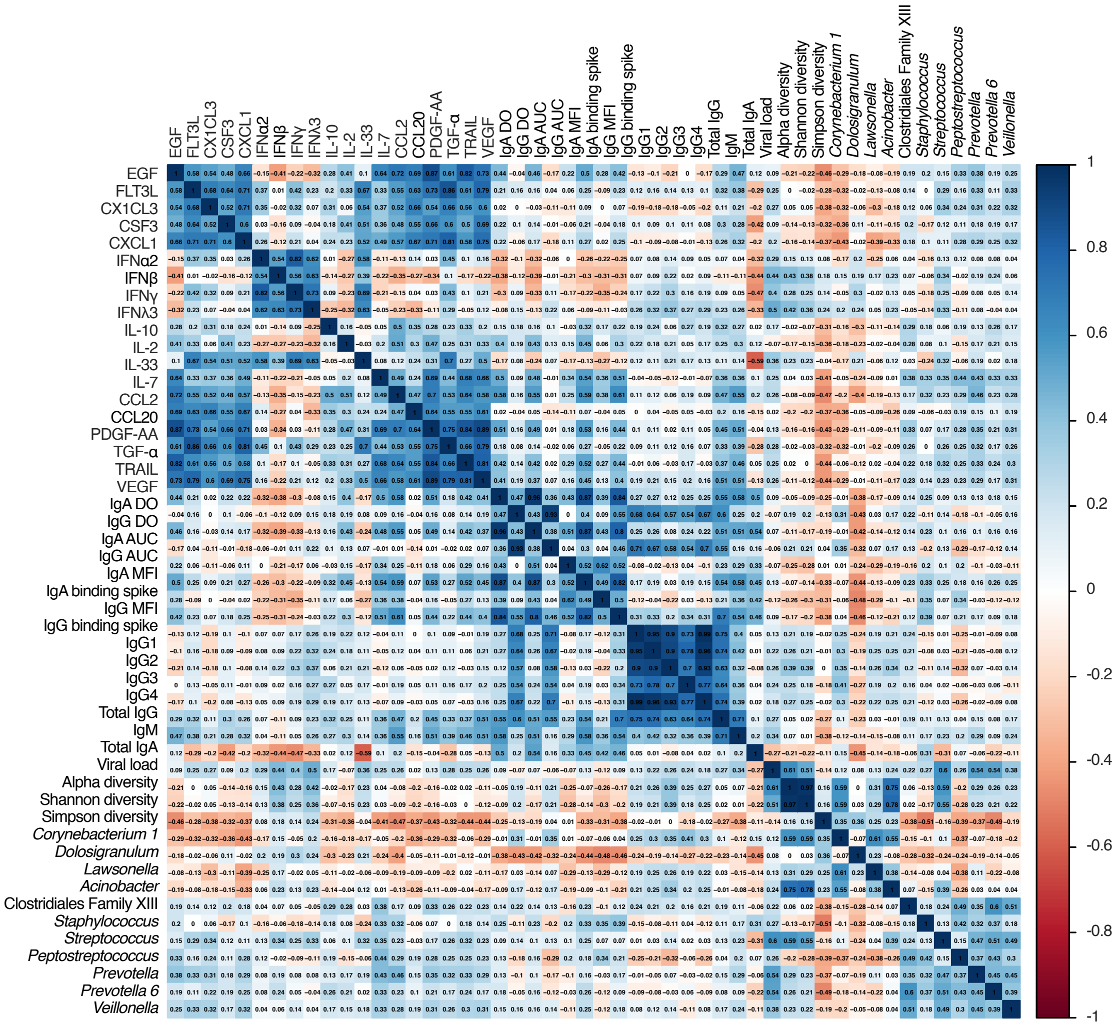
